# PrEgabalin for Treatment Resistant generalised Anxiety disorder

**DOI:** 10.64898/2026.09.01.26359217

**Authors:** Glyn Lewis, Nick Freemantle, Hakim-Moulay Dehbi, Charlotte Clarke

## Abstract

This document describes the Statistical Analysis Plan (SAP) for PETRA, a randomised controlled trial in people with generalised anxiety disorder comparing pregabalin plus an antidepressant and standard care, with placebo plus an antidepressant and standard care, with respect to the primary outcome of the GAD-7 score at week 12.

## 1 INTRODUCTION

### 1.1 Background and rationale

Generalised anxiety disorder (GAD) is characterised by at least six months of symptoms including disproportionate worry, nervousness, poor concentration and sleep disturbance. The prevalence of GAD is 6%, higher than depression (3%) in the UK ^1^ but is less likely to present and be recognised in primary care. However, over the past few years the rate of GAD recorded in primary care has increased by 50% and especially in younger, working-age adults aged 18-35 years, in whom rates of presentation have more than doubled over the past decade ^2^.

Reducing GAD symptoms could also lead to benefit for other comorbid conditions. For example, generalised anxiety can increase later depressive symptoms so treatments that improve anxiety symptoms could also reduce depressive symptoms ^3^.

Psychological treatments (e.g. cognitive behavioural therapy) and antidepressants are the main evidence-based options for GAD ^4^. The evidence so far is that psychological and pharmacological benefits are additive for most depressive and anxiety disorders ^5^ optimising pharmacological treatment will also be of benefit for those receiving psychological treatments. Access to psychological treatments is limited, with lengthening waiting, so research into improving the pharmacological treatment of anxiety is timely.

There is evidence that selective serotonin reuptake inhibitors (SSRIs), serotonin noradrenaline reuptake inhibitors (SNRIs) and mirtazapine (a noradrenergic and specific serotonergic antidepressant) are effective for people with GAD. After antidepressant treatment, about 50% of people still have significant generalised anxiety ^6 7^, even if there has been some improvement. There is currently no consensus and much clinical uncertainty about what pharmacological treatments should be used after non-response or partial response to antidepressants. We are only aware of one study that has investigated pregabalin as an adjunctive treatment in primary care ^8^ and this supports its use, but the treatment effect was relatively small and of unknown clinical importance. Furthermore, the study gave little information on how participants were recruited and their clinical characteristics so the generalisability to the NHS or other health care settings is uncertain.

### 1.2 Objectives

To investigate whether pregabalin compared to placebo, in addition to an antidepressant, is an effective and cost-effective treatment for generalised anxiety disorder in people who have not responded to antidepressant treatment.

To investigate any adverse effects associated with combined treatment of pregabalin and antidepressants.

To investigate withdrawal symptoms from pregabalin when it is used in combination with antidepressants.

To investigate the acceptability of prescribing pregabalin in addition to antidepressants for GAD from the perspectives of patients and general practitioners using qualitative methods.

### 1.3 Estimand framework (primary outcome)

**Table 1:** Estimand framework as it applies to the primary outcome.

| Characteristic of estimand | Definition and method of analysis |
| --- | --- |
| Population | Participants who have a diagnosis of generalised anxiety disorder, who have not responded to at least two antidepressant medications, who are still currently taking antidepressant medication, who are in the community and who do not fulfil exclusion criteria. |
| Treatment conditions | Pregabalin medication versus placebo in addition to care as usual |
| Primary outcome | Score on the Generalised Anxiety Disorder scale (GAD-7) at 12 weeks post randomisation |
| Summary measure | Ratio of geometric means of the score on the Generalised Anxiety Disorder scale (GAD-7) at 12 weeks post randomisation between those randomised to placebo versus those randomised to pregabalin (adjusting for baseline scores). |
| Intercurrent Events |  |
| Death before outcome | While alive policy |
| Withdrawal from randomised medication (stays in trial) | Treatment policy |
| Increased use of alcohol | Treatment policy |
| Psychiatric admission | Treatment policy |
| Commencement of psychological therapies | Treatment policy |

## 2 STUDY METHODS

### 2.1 Trial design

A double-blind randomised controlled trial for people with generalised anxiety disorder, who have been treated with at least two antidepressants, one of which failed, and who are currently still taking the other antidepressant. Participants will be randomised with a 1:1 ratio, to pregabalin or placebo with both groups also receiving usual care from their general practitioner who will continue to prescribe their existing antidepressant.

### 2.2 Randomisation

Randomisation will be by minimisation using a 70%-30% biased coin, so that the overall number of participants will asymptote to 1:1 over time. Minimisation factors are site (4 sites), GAD-7 score at baseline dichotomised (≤11/>11), and type of antidepressant (SSRI/other).

### 2.3 Sample size

We estimated the minimal clinically important difference for the GAD-7 as about a 20% reduction ^9^. From the Cobalt and MiR trial results at 6 months our MCID estimate is 1.6 or 1.9 GAD-7 points and standard deviations (SDs) were 5.6 or 5.8. In those trials the correlation between baseline and 12-week GAD7 scores was 0.47 so this leads to an effective SD of 4.9. For a difference of 1.6 GAD-7 points we need 199 per group for 90% power at the 5% significance level. Our target is 498 patients allowing for 20% attrition or other methodological challenges at 12 weeks, though we would expect to achieve substantially better follow up. For the withdrawal study, the power depends upon the position on the binomial distribution, but clinically important differences in withdrawal effects will be detectable. For example, we will have 85% power to find a difference between the groups where the control condition has a proportion of 2.5% of withdrawal symptoms and the pregabalin group 10%.

### 2.4 Framework

PETRA is a superiority trial. The primary purpose of which is to assess whether pregabalin, compared to placebo, plus antidepressant is effective at reducing generalised anxiety disorder in people who have not responded to antidepressant treatment alone.

### 2.5 Statistical interim analyses and stopping guidance

There are no planned interim analyses. Monitoring of the safety of the trial will be undertaken by the Data Monitoring and Ethics Committee (DMEC) which will have untrammelled access to the trial data, and whose work is governed by a separate charter.

### 2.6 Timing of final analysis

The final analysis will start when all data has been entered into the database, all corresponding queries have been resolved, and the database has been locked.

### 2.7 Timing of outcome assessments

The timing of outcome assessments is provided in section 5.2 (Participant timeline) of the protocol. These are 3, 6, 12, 26 and ∼30 weeks post baseline.

The primary outcome is at 12 weeks.

## 3 STATISTICAL PRINCIPLES

### 3.1 Confidence intervals and p-values

All confidence intervals presented will be 95% and two-sided, unless otherwise specified.

### 3.2 Analysis population and Missing Data

The primary outcome analysis will be conducted following the intention-to-treat (ITT) principle where all randomised participants are analysed in their allocated treatment group regardless of whether they receive their randomised treatment. However, those who never ingest the IMP and return the unopened package to the research team will be excluded from the full analysis set and considered non-randomised. Additionally, on a case-by-case basis, a participant may be excluded from the full analysis set if they fail (retrospectively) to satisfy one of the major entry criteria as detailed in ICH E9 section 5.2.1.

Missing baseline data are not anticipated since baseline data must be recorded to allocate treatment. All participants with reported outcome data within the time window defined in the protocol will be included in the analysis. All efforts will be made to ensure that the primary outcome data is collected for all patients at 12 weeks. Should there be substantial amounts of missing data, we will consider further analyses to examine the effect that missing data may have on our findings. We will conduct descriptive and regression-based analyses to examine potential predictors of missing outcome data, in order to assess potential missing data mechanisms.

An ITT analysis of all patients with reported outcome data will be performed for all secondary outcomes.

## 4 TRIAL POPULATION

### 4.1 Screening, recruitment, withdrawal/ follow-up

Participant flow from those screened through to enrolment, follow-up, and inclusion in the final analysis, as well as those who withdraw or are lost to follow-up, will be summarised in a CONSORT ^10^ flowchart constructed by the Trial Manager with input from the Trial Statistician.

### 4.2 Eligibility

Eligibility and inclusion/ exclusion criteria are provided in sections 3.1 and 3.2 of the protocol.

### 4.3 Baseline patient characteristics

Baseline characteristics will be summarised for all randomised participants in the trial. Summary measures for the baseline characteristics will be presented as mean and standard deviation for continuous (approximately) normally distributed variables, medians and interquartile ranges for non-normally distributed continuous variables, and frequencies and percentages for categorical variables. We will plot histograms of continuous variables to assess normality. These analyses will be done overall and by randomised group.

## 5 ANALYSIS

### 5.1 Outcome definitions

#### 5.1.1 Primary Outcome

The primary outcome for this trial is the GAD-7 ^11^ at 12 weeks post randomisation. This is a measure of generalised anxiety symptoms. There are seven items which are scored 0 (not at all) to 3 (nearly every day). Items are summed to give a score between 0 and 21, with a higher score indicating more severe anxiety symptoms. Given that GAD-7 is known to be right skewed, the natural logarithm will be used in the statistical model.

If anyone scores 0, a small amount (e.g. 0.1) will be added to the score before taking natural logs to enable everyone with data to be included in analysis. If there are one or two items missing from a participant’s questionnaire, items will be replaced by the mean of the items present. If there are more than two items missing, the questionnaire will be considered missing for that participant.

#### 5.1.2 Secondary Outcomes

- Natural log and total score of GAD-7 at weeks 3, 6, 26, and 30.
- Presence of anxiety (presence or absence based on a cut-off of GAD-7 ≥10) at weeks 3, 6, 12, 26, and 30.
- Natural log and total score of Patient Health Questionnaire 9 (PHQ-9) ^12^ at weeks 3, 6, 12, 26, and 30.
- Suicidal thinking from PHQ9 at weeks 3, 6, 12, 26, and 30.
- Panic symptoms based on the Patient Health Questionnaire for Panic Disorder (PHQ-PD) ^13^ at weeks 3, 6, 12, 26, and 30.
- Generic measure of health-related quality of life using questions from the EQ-5D-5L questions at weeks 3, 6, 12, 26, and 30.
- Modified SF-36 Health Survey mental component ^14^ at weeks 3, 6, 12, 26, and 30.
- Modified SF-36 Health Survey physical component ^14^ at weeks 3, 6, 12, 26, and 30.
- Self-reported global improvement score ^15^ at weeks 3, 6, 12, 26, and 30.
- Adherence to study medication at weeks 3, 6, 12, and 26.
- Continuation of antidepressant use at weeks 3, 6, 12, 26, and 30
- Safety and tolerability of Pregabalin as indicated by changes in Toronto scale total score for antidepressants ^16^ and waist circumference, new or increased thoughts of self-harm/suicide, and pregabalin withdrawal symptoms.
- Alcohol consumption based on the AUDIT PC score at weeks 3, 6, 12, 26, and 30.
- Benzodiazepine use at week 3, 6, 12, 26, and 30.
- Whether participants thought themselves to be on the active drug or placebo at weeks 3, 6, 12, and 26.

### 5.2 Analysis Methods

The results of the analyses will be reported following the principle of the ICH E3 guidelines on the Structure and Content of Clinical Study Reports ^2^. All statistical analysis will be performed using R version 4.4.1 or higher.

Data at all follow-up time points will be summarised. Continuous variables will be reported using mean (SD) or median (IQR) depending on distribution. Categorical variables will be reported using frequency (%).

We will not adjust for multiple testing. The primary outcome will be analysed at a two-sided significance level of 0.05. Analyses of all other outcomes will be reported with nominal p-values.

#### 5.2.1 Adjustment factors

All models will be adjusted for the minimisation factors, unless stated otherwise.

- Site: Bristol/Keele/Liverpool/UCL
- Baseline GAD-7 score: ≤11 or >11
- Class of antidepressant participants are taking at baseline: SSRI or other

#### 5.2.2 Primary Outcome Analysis

A mixed-effect linear model will be used to estimate the difference in log GAD-7 between the intervention groups at 12 weeks post-randomisation.

The model will include fixed effects for intervention group (pregabalin or placebo), time (baseline and 12 weeks). The minimisation factors (site and class of concomitant antidepressant) will be included as covariates. The minimisation factor related to GAD-7 will not be included because the baseline GAD-7 is part of the dependent variable in the model. A random effect for patient will be included to take account for clustering. This approach is analogous to the data analysis method used in ANTLER ^17^.

For interpretation purposes, the coefficient associated with the primary outcome, as well as the confidence interval (CI), will be exponentiated. It will be presented as ratio of geometric means (RoGM) between the groups with 95% CI.

##### Sensitivity analysis of primary outcome

The following sensitivity analyses on the primary outcome to assess the robustness of results will be considered:

- If there are more than 10% missing data on the primary outcome at 12 weeks:
◦ we will explore baseline predictors of missing GAD-7 at 12 weeks using logistic regression. Under a missing-at-random assumption, we will perform multiple imputation of missing 12-week GAD-7 values using a model including baseline GAD-7, treatment allocation, site, antidepressant class, and other baseline covariates considered clinically relevant. The primary analysis will be re-fitted in each imputed dataset.
◦ the primary analysis will be repeated with additional adjustments for any baseline variables that show notable imbalance between treatment groups to assess whether chance imbalance impacts the estimated treatment effect.
- Depending on the observed levels of adherence to medication during the primary time frame up to 12 weeks, a Complier Average Causal Effect (CACE) will be considered.

#### 5.2.3 Secondary outcome analysis

##### Continuous Secondary Outcomes

Each of the following continuous secondary outcome measures will be analysed using a separate linear mixed-effect model. The parameterisation will be the same as the model of the primary outcome:

- Natural log and total score of GAD-7 at weeks 3, 6, 12 and 26
- Natural log and total score of PHQ9 at weeks 3, 6, 12, and 26
- Modified SF-36 Health Survey at weeks 3, 6, 12, and 26.

For GAD-7 outcomes, we will not include the GAD-7 minimisation factor as baseline GAD-7 score will be used as dependent variable in the linear mixed model. For all other outcomes, we will include all minimisation factors.

We will include time as a continuous variable in the mixed model. The mean at the various time points will be provided according to this modelling strategy. We will also model outcomes using time as a categorical variable. We will provide estimates from this approach for each time point with 95% confidence intervals.

Similarly to the primary outcome, for interpretation purposes we will exponentiate coefficients and CIs to provide the RoGM between groups.

##### Categorical Secondary Outcomes

Each of the following categorical secondary outcome measures will be analysed using a mixed-effect logistic regression model including all follow-up time points (weeks 3, 6, 12, and 26) and adjusted for minimisation factors:

- Dichotomised total GAD-7 based on the cut off ≥10 vs. <10 at weeks 3, 6, 12, and 26.
- Presence of panic symptoms based on the PHQ-PD at week 3, 6, 12, and 26 (based on the answer to all 5 questions pertaining to anxiety attacks being answered yes (i.e. a score of 5 vs ≤4), as well as based on the answer only to the first question (“In the last 4 weeks, have you had an anxiety attack – suddenly feeling fear or panic?”) being answered yes).
- Self-reported global improvement score at weeks 3, 6, 12, and 26.
- AUDIT PC (dichotomised as score ≥5 vs <5) at week 3, 6, 12, and 26.
- Adherence to medication at week 3, 6, 12, and 26
- Continuation of antidepressant use at week 3, 6, 12, and 26.
- Benzodiazepine use at week 3, 6, 12, and 26
- Thoughts of self-harm / suicide over previous 2 weeks at week 3, 6, 12, and 26 based on PHQ-9 question 9 (dichotomised as score 0 vs 1/2/3, and as score 1/2 vs 3/4)
- Whether participants think they are on the active drug or placebo at week 3, 6, 12, and 26

In addition to logistic regressions, these outcomes will be summarised descriptively using counts and proportions, by randomisation group and time point (weeks 3, 6, 12, 26).

##### Adverse events and serious adverse events

Changes in waist circumference, and Toronto scale total score for antidepressants at weeks 3, 6, 12, and 26, will be summarised by arms using descriptive statistics, and compared using t-test (or non-parametric equivalent).

The proportion of patients experiencing at least one adverse event and those experiencing at least one serious adverse event will be summarised by treatment arm. Chi-square tests will be used to compare study arms. The number and percentage of adverse events and serious adverse events will be presented descriptively by arm. Information on grades of events and whether the events are expected or unexpected will be presented.

The number of participants withdrawing from the trial due to an AE or SAE will be summarised by treatment group.

##### Analysis of the tapering phase

After the week 26 assessment, participants will be instructed to taper their medication so that they will no longer be taking medication at the 30-week time point. The main outcome of interest in this part of the study is to examine the withdrawal symptoms that emerge during tapering. Given the lack of existing knowledge about withdrawal symptoms, we will use the total score of the adverse effects scale (see above) as the main outcome. We will also examine the GAD7 and PHQ9 scores at 30 weeks as these would also be expected to change following tapering of pregabalin.

The analysis will include the participants who have follow up data at the 26-week time point. This group is expected to be substantially smaller than the original randomised sample. We will therefore compare the baseline characteristics of the two randomised groups who have data at 26 weeks and adjust the analysis for baseline factors imbalanced or associated with missing data at 26 weeks. Our analysis of the Toronto scale total score, GAD7 and PHQ9 data will use a linear mixed model similar to that proposed for the primary outcome.

#### 5.2.4 Subgroup analyses

We will calculate interactions between randomised groups and each subgroup variable (below) with respect to the primary outcome. Other variables in these models will be the same as those included in the main analyses of the primary (GAD7 at week 12). We will present the p-value for interaction. The subgroups investigated will be:

- Class of antidepressant medication at baseline (SSRI vs other)
- Baseline anxiety score (dichotomised both using GAD-7 ≤11/>11 and at the median baseline GAD-7 total score)
- Baseline depression score (dichotomised at the median PHQ-9 total score)

Following the interaction analysis, we will carry out analyses by the subgroups listed above. We will present coefficients or odds ratios as appropriate and 95% CI for the randomisation variable.

#### 5.2.5 Safety reporting

As pregabalin is a licensed drug, we are only collecting data on AEs of interest via our CRFs, as described above as a secondary outcome. SAEs will be reported separately by randomised group. SAEs, SARs and SUSARs reported during the treatment period will be presented by treatment arm and grade. Full listings of safety events will be provided in statistical reporting.

## 6 ECONOMIC EVALUATION

The aim of the economic evaluation is to estimate the incremental cost per quality-adjusted life year (QALY) gained for Pregabalin, usual care and their existing antidepressant compared to their existing antidepressant and usual care only from their GP over 26 weeks from an NHS health and social care perspective.

Primary analysis will be within-trial cost-utility analysis over 26 weeks with outcome measured as QALYs calculated using health related quality of life questions form the EQ-5D-5L (2026 value set). Secondary analyses will include: (i) using the EQ-5D-5L Crosswalk Index Value Calculator to estimate the index values; (ii) using the SF-12 and the SF-6D algorithm to calculate QALYs; and (iii) including the cost of productivity loss.

### 6.1 Outcomes

The following outcomes will be used for the economic evaluation:

- Quality of life using the five health related quality of life questions from EQ-5D-5L questionnaire. This is a five item, five level questionnaire, scored 1 (no problem) to 5 (extreme problems). The questionnaire is completed at baseline, 3, 6, 12, 26 and 30 weeks follow-up. The new EQ-5D-5L value set will be used to estimate index values^18^. For a secondary analysis, the EQ-5D-5L Crosswalk Index Value Calculator will be used to estimate the index values^19^. The index values will be used in the QALY calculation. QALYs will be calculated as the area under the curve adjusting for baseline utility^20^. We will analyse the difference in QALYs between the intervention and the placebo arm using linear regression with covariates for site (4 sites), GAD-7 score at baseline dichotomised (≤11/>11), and type of antidepressant (SSRI/other). The confidence intervals will be obtained using bootstrapping procedure with 1,000 bootstrap samples.
- Modified SF-36 Health Survey. It is a patient-reported generic measure of health-related quality of life. It consists of 8 dimensions that in total include 35 multi-level items. Reduced version of the SF-36 was derived and valued using standard gamble method to create a value set (SF-6D)^21^. We will use the responses from the SF-36 to calculate index values that will be used in the QALY calculation. We will calculate the area under the curve adjusting for baseline and estimate the difference in QALYs between the intervention and the placebo arm using linear regression controlling for site (4 sites), GAD-7 score at baseline dichotomised (≤11/>11), and type of antidepressant (SSRI/other). The confidence intervals will be obtained using bootstrapping procedure with 1,000 bootstrap samples.
- Healthcare resource use will be collected from GP records and self-reported bespoke questionnaires at baseline and 26 weeks follow-up.

### 6.2 Cost data

Cost data are comprised of cost of the intervention, cost of healthcare resource use and cost of absenteeism.

#### Cost of the intervention

Information collected on the intervention medicine (pregabalin) in the CRFs will be costed using the British National Formulary (BNF)^22^. We will report mean cost per participant of pregabalin.

#### Cost of healthcare resource use

Information on healthcare resource use will be collected from GP records and self-reported bespoke questionnaires. The bespoke, self-completed health and social care resource use questionnaire collected at baseline and 26 weeks follow-up asking about the previous 6-months.

From GP records we will obtain the following resource use:

- Primary care consultations for the patient in the 6 months prior to randomisation
- Primary care consultations for the patient in the 6 months after randomisation
- Other antidepressant medication prescriptions

The self-reported questionnaires will collect data on:

- talking therapies
- other health care professionals (mental health nurse, occupational therapist, social worker, clinical psychologist)
- active relaxation
- other community-based and emergency care (NHS walk-in centres, ambulance, 111, A&E attendance etc)
- hospital care (outpatient contacts)
- hospital care (inpatient stays)

Descriptive statistics for the percentage of participants and mean number of contacts for each type of resource will be reported at baseline and 26 weeks follow-up. Information on data completeness will also be reported. The cost components will be costed for each participant using unit costs from the most recent Unit Costs of Health and Social Care published by the Personal Social Services Research Unit (PSSRU)^23^ and the NHS Reference costs^24^. Mean cost per participant and standard deviation for the intervention arm versus control arm will be reported by type of service use at baseline and 26 weeks follow-up.

Other antidepressant medication use will be collected from GP files and costed using the BNF. We will calculate the mean cost per participant of other antidepressant medications. Mean incremental cost per patient of the intervention compared to control and 95% confidence intervals will be calculated based on 95% bias corrected and adjusted bootstrap replications adjusting for baseline resource use and with covariates for site (4 sites), GAD-7 score at baseline dichotomised (≤11/>11), and type of antidepressant (SSRI/other).

A sensitivity analysis will calculate the difference in costs between intervention and control arm using a generalised linear model (GLM). This directly models both the mean and variance functions on the original scale of cost. The modified Parks test will be used to identify the family for GLM. The model will include covariates for site (4 sites), GAD-7 score at baseline dichotomised (≤11/>11), and type of antidepressant (SSRI/other). We will report 95% confidence intervals.

#### Total costs

The overall mean cost per participant will be reported and will be the sum of the costs described above. For participants who die during the follow-up, their recorded costs will be included up to their date of death. We will analyse the difference in total costs between intervention and control arm using a bias corrected and adjusted bootstrapping as described above.

#### Cost of productivity loss

The cost of lost employment for the intervention arm versus control arm will be calculated from participants completed bespoke questionnaire at baseline and 26 weeks follow-up using the human capital approach. The approach is to multiply the number of lost hours by the median hourly wage based on sex, occupation and full-time/part-time employment in the United Kingdom (UK)^25^.

### 6.3 Primary analysis

We will use seemingly unrelated regression to report the incremental mean difference in costs and QALYs adjusting for baseline HRQL and costs. We will report point estimates and 95% confidence intervals. The point estimates will be used to calculate the incremental cost-effectiveness ratio (ICER).

Uncertainty in the point estimate of cost per QALY will be quantified using bootstrapping methods to calculate confidence intervals around the ICER.

The bootstrap results will be used to generate the cost-effectiveness acceptability curve (CEAC)^26^: the probability that pregabalin is cost-effective compared to usual care at 26 weeks for a range of willingness-to-pay for an additional QALY. A cost-effectiveness plane (CEP) of the bootstrap results will also be reported.

Analyses will be performed using STATA programs (* .do).

### 6.4 Secondary analyses

We will conduct a series of secondary analyses within-trial analyses:

- Cost-utility analysis using EQ-5D-5L index values estimated using the Crosswalk Index Value Calculator
- Cost-utility analysis using SF-6D index values
- Cost-utility analysis using wider perspective (including cost of productivity loss)

The following subgroup analyses will be conducted:

- class of antidepressant medication at baseline,
- baseline anxiety score,
- baseline depression score.

### 6.5 Missing data

In line with the SAP, the primary analysis will be conducted following the intention-to-treat principle in accordance with the randomised intervention.

The primary analysis will be a complete case analysis unless >15% of participants are missing an ICER. If >15% of participants are missing an ICER, we will examine the data for predictors of missingness assuming that data are missing at random. Initial analyses will include mixed model (LMM) to account for missing data as recommended in Gabrio et al.^27^ with more complex multiple imputation models for health economic outcomes being considered in a stepped approach as recommended by Faria et al.^28^

## Data Availability

No datasets were generated or analysed during the current study. This manuscript describes the pre-specified statistical analysis plan for the PETRA trial.

## 8 REVISION HISTORY

Version 0.1: prepared by Louise Marston

Version 0.2: amended by Charlotte Clarke and Hakim-Moulay Dehbi. Changes were made to reflect discussions with TSC.

Version 0.3: refinement of outcome definition based on conversations with CI Professor Glyn Lewis

Version 0.4: further refinement of analysis methods for continuous outcomes

Version 0.5: further refinement of secondary outcomes and addition of health economics evaluation

Version 2.0: Added an abstract required for submission to the preprint Medrxiv

## 9 APPENDICES

## Notes

### Competing Interest Statement

The authors have declared no competing interest.

### Clinical Trial

ISRCTN16993990

### Author Declarations

The East Midlands - Leicester Central Research Ethics Committee of the Health Research Authority gave ethical approval for this work (REC reference: 23/EM/0192).

### Funding

- National Institute for Health and Care Research, Health Technology Assessment (NIHR HTA): 134074

